# Factors Associated with Long Covid Symptoms in an Online Cohort Study

**DOI:** 10.1101/2022.12.01.22282987

**Authors:** Matthew S. Durstenfeld, Michael J. Peluso, Noah D. Peyser, Feng Lin, Sara J. Knight, Audrey Djibo, Rasha Khatib, Heather Kitzman, Emily O’Brien, Natasha Williams, Carmen Isasi, John Kornak, Thomas W. Carton, Jeffrey E. Olgin, Mark J. Pletcher, Gregory M. Marcus, Alexis L. Beatty

## Abstract

**Importance:** Prolonged symptoms following SARS-CoV-2 infection, or Long COVID, is common, but few prospective studies of Long COVID risk factors have been conducted.

**Objective:** To determine whether sociodemographic factors, lifestyle, or medical history preceding COVID-19 or characteristics of acute SARS-CoV-2 infection are associated with Long COVID.

**Design:** Cohort study with longitudinal assessment of symptoms before, during, and after SARS-CoV-2 infection, and cross-sectional assessment of Long COVID symptoms using data from the COVID-19 Citizen Science (CCS) study.

**Setting:** CCS is an online cohort study that began enrolling March 26, 2020. We included data collected between March 26, 2020, and May 18, 2022.

**Participants:** Adult CCS participants who reported a positive SARS-CoV-2 test result (PCR, Antigen, or Antibody) more than 30 days prior to May 4, 2022, were surveyed.

**Exposures:** Age, sex, race/ethnicity, education, employment, socioeconomic status/financial insecurity, self-reported medical history, vaccination status, time of infection (variant wave), number of acute symptoms, pre-COVID depression, anxiety, alcohol and drug use, sleep, exercise.

**Main Outcome:** Presence of at least 1 Long COVID symptom greater than 1 month after acute infection. Sensitivity analyses were performed considering only symptoms beyond 3 months and only severe symptoms.

**Results:** 13,305 participants reported a SARS-CoV-2 positive test more than 30 days prior, 1480 (11.1% of eligible) responded to a survey about Long COVID symptoms, and 476 (32.2% of respondents) reported Long COVID symptoms (median 360 days after infection).

Respondents’ mean age was 53 and 1017 (69%) were female. Common Long COVID symptoms included fatigue, reported by 230/476 (48.3%), shortness of breath (109, 22.9%), confusion/brain fog (108, 22.7%), headache (103, 21.6%), and altered taste or smell (98, 20.6%). In multivariable models, number of acute COVID-19 symptoms (OR 1.30 per symptom, 95%CI 1.20-1.40), lower socioeconomic status/financial insecurity (OR 1.62, 95%CI 1.02-2.63), pre-infection depression (OR 1.08, 95%CI 1.01-1.16), and earlier variants (OR 0.37 for Omicron compared to ancestral strain, 95%CI 0.15-0.90) were associated with Long COVID symptoms.

**Conclusions and Relevance:** Variant wave, severity of acute infection, lower socioeconomic status and pre-existing depression are associated with Long COVID symptoms.

**Key Points:** *Question:* What are the patterns of symptoms and risk factors for Long COVID among SARS-CoV-2 infected individuals? Findings
Persistent symptoms were highly prevalent, especially fatigue, shortness of breath, headache, brain fog/confusion, and altered taste/smell, which persisted beyond 1 year among 56% of participants with symptoms; a minority of participants reported severe Long COVID symptoms. Number of acute symptoms during acute SARS-CoV-2 infection, financial insecurity, pre-existing depression, and infection with earlier variants are associated with prevalent Long COVID symptoms independent of vaccination, medical history, and other factors.

Meaning
Severity of acute infection, SARS-CoV-2 variant, and financial insecurity and depression are associated with Long COVID symptoms.

## Introduction

Symptoms attributable to COVID-19, including fatigue, memory/concentration problems (“brain fog”), and shortness of breath may persist after acute infection. Symptoms may be due to Long COVID, a type of post-acute sequelae of COVID-19 (PASC) not explainable by other known medical conditions.^1^ While prevalence estimates vary, Long COVID may occur in up to 30% of individuals following SARS-CoV-2 infection.^2-7^ Most individuals with Long COVID were not hospitalized for acute infection, and Long COVID can occur regardless of illness severity, vaccination status, and SARS-CoV-2-targeted treatment,^8,9^ although the risk may be lower among vaccinated individuals and those with asymptomatic infection.^9,10^ Symptoms have been reported for up to 24 months,^11^ and there are currently no proven treatments for Long COVID.

The existing literature on risk factors for Long COVID largely relies on assessments performed after SARS-CoV-2 infection, captured in electronic health record diagnostic codes, or focused on individuals recruited from Long COVID clinics or after hospitalization for acute COVID-19.^12-14^ Furthermore, cohort studies which require in-person participation may exclude individuals unable to attend study visits or far from research sites, and thus survey-based approaches, particularly those which leverage pre-infection data, may contribute to our understanding of Long COVID and its antecedent factors.^10,15,16^ The objectives of this study were to estimate Long COVID symptom prevalence and determine whether sociodemographic factors, lifestyle, or medical history preceding COVID-19, or characteristics of acute SARS-CoV-2 infection were associated with development of Long COVID symptoms.

## Methods

### Design, Setting, and Participants

The COVID-19 Citizen Science (CCS) study is an online cohort study that began enrolling participants on March 26, 2020.^17^ CCS is hosted on the Eureka Research Platform (University of California, San Francisco), a digital platform for clinical research studies including a mobile application (app) and web-based software. Participants are recruited through email invitations to participants in other Eureka Research Platform studies, press releases, word-of-mouth, and by partner organizations. Participants must be 18 years of age or older, register for a Eureka Research Platform account, have an iOS or Android smartphone with a cell phone number (or enroll in the web-based study launched January 21, 2021), agree to participate in English, and provide consent. There were no geographic restrictions, but 98% were United States residents. After providing electronic consent, participants complete baseline, daily, weekly, and monthly surveys. CCS methods have been previously described.^17^ For this analysis, we included data collected March 26, 2020 to May 18, 2022. The primary analyses included all individuals who reported a positive COVID-19 test result (PCR, Antigen, or Antibody) more than 30 days prior to May 4, 2022 and responded to a survey about Long COVID symptoms, but for longitudinal symptom comparison we also included those without a positive COVID-19 test and compared to those without SARS-CoV-2 infection. Sample size was not determined a priori. The study was reviewed and approved by the UCSF Institutional Review Board (#17-21879). Results are reported in accordance with STROBE guidelines.^18^

### Long COVID Symptoms

Participants who reported a COVID-19 positive test more than 30 days prior were offered a cross-sectional survey about Long COVID Symptoms in January 2022 and May 2022 (surveys went out on different days depending on participant’s last answered survey). The survey asked about the presence, duration, and severity of Long COVID symptoms using a non-validated instrument. Severity was assessed using a Likert-scale asked for each reported symptom: “How bad do you think these symptoms were?” (1-5, very mild to very severe). Additionally, the survey asked about health care use and missed days of work or school due to Long COVID symptoms.

### Longitudinal Symptoms

In addition to cross-sectional surveys, participants were surveyed on a daily or weekly basis regarding symptoms including: a scratchy throat, a painful sore throat, a cough, a runny nose, symptoms of fever or chills, a temperature >100.4°F or 38.0°C, muscle aches, nausea, vomiting, or diarrhea, shortness of breath, unable to taste or smell, and red or painful eyes. We did not use these surveys to classify people as having Long COVID. We categorized the surveys for each individual by time period relative to date of SARS-CoV-2 positive test into 30-60 days pre-infection, 0-30 days pre-infection, 0-30 days post-infection, 30-90 days post-infection, 90-180 days post infection, 180-365 days post infection, and >365 days post infection. For each period, we summed the proportion of respondents averaging _≥_1 symptom and the average number of symptoms reported by Long COVID status among respondents to the cross-sectional survey and among SARS-CoV-2 infected non-respondents, and uninfected individuals (averaged over all time points since March 2020).

### Other variables

Variant wave was classified by the date of first positive test: Initial (prior to March 11, 2021), Alpha (March 11, 2021-July 3 2021), Delta (July 4, 2021-December 25 2021), and Omicron (December 26, 2021-April 4, 2022).^19^ Participants self-reported demographics, medical history, and vaccine history via surveys. Most participants (969/1480, 65.5%) completed baseline surveys prior to SARS-CoV-2 infection, while the remainder enrolled after infection. Participants were queried about lifestyle factors before and after COVID including number of days per week they exercised, typical amount of sleep, and number of alcoholic drinks they consumed.

Standardized instruments including the PHQ-8 for depression^20^ and GAD-7 for anxiety^21^ were used in a self-administered format and were assessed before and after COVID for those who completed surveys prior to infection. Socioeconomic status was assessed using the MacArthur Scale of Subjective Social Status.^22^

### Statistical Analysis

The presence of Long COVID symptoms was defined based _≥_1 symptom reported more than 30 days after SARS-CoV-2 positive test on the cross-sectional survey. First, we compared demographics, pre-COVID medical history, socioeconomic variables, and lifestyle patterns among those with Long COVID symptoms compared to those without Long COVID symptoms using Chi-squared tests for categorical variables and T-tests for continuous variables. Then we described the prevalence of each reported symptom and patterns of symptom persistence. To assess risk factors associated with Long COVID and adjust for potential confounders, we constructed multivariable logistic regression models in a pre-specified staged approach using a complete case approach to missing data. In an initial (baseline) model (Stage 1) we included age, sex, variant wave, number of symptoms during acute infection, and past medical history. For the next model (Stage 2) we added vaccine status and timing, Hispanic ethnicity, and sociodemographic variables including socioeconomic status, education level, and employment in healthcare. In the final model (Stage 3), in which we prespecified including pre-COVID variables with p<0.10 in univariate analysis, we added pre-COVID anxiety, depression, and financial insecurity. We additionally conducted sensitivity analyses considering only those with pre-COVID baseline assessments, only those with persistent symptoms and only those with severe/very severe symptoms. Statistical significance was considered to be p<0.05 for all analyses other than the prespecified potential predictor selection process in Stage 3. All analyses were conducted with SAS version 9.4 (Cary, NC).

## Results

As of May 18, 2022, 13305 participants reported a diagnosis of COVID-19 more than 30 days prior to the survey, 1480 (11.1%) responded to a survey about Long COVID symptoms, and 476 of these (32.2% of respondents) reported Long COVID symptoms. Compared to non-respondents, survey respondents were more likely to be infected during the Omicron wave and less likely during the Initial wave, more likely to have been vaccinated prior to infection, and had a higher number of acute symptoms (p<0.001 for each). Among those with Long COVID symptoms, mean age was 53.1 (standard deviation 13.3) and 356 (75.1%) female (Table 1). Among those with Long COVID symptoms, the median time from first SARS-CoV-2 positive test was 360 days (IQR 129, 506) and among those without was 129 days (IQR 108, 343).

**Table 1.** Participant Characteristics Baseline and post-COVID characteristics among those with Long COVID symptoms, without Long COVID symptoms, and non-responders. P values are for univariate unadjusted comparison between those with and without Long COVID symptoms reported on the cross-sectional survey using Chi-squared tests for categorical variables and T-tests for continuous variables. ^a^Participants could check all of the above so totals do not add to 100% and each race/ethnicity was considered separately rather than as a single categorical variable.

|  | Long COVID Symptoms, N = 476 | No Long COVID Symptoms, N = 1004 | Didn't answer survey, N = 11825 | P-value comparing symptoms vs. no symptoms |
| --- | --- | --- | --- | --- |
| Age, mean (SD) | 53.13 (13.27) | 52.50 (14.13) |  | 0.42 |
| Female sex at birth | 356 (75.11%) | 661 (65.90%) | 8153 (69.34%) | 0.0004 |
| Race/Ethnicity<sup>a</sup> |  |  |  |  |
| White | 433 (92.52%) | 924 (92.77%) | 10374 (89.12%) | 0.86 |
| Black or African American | 15 (3.21%) | 33 (3.31%) | 493 (4.24%) | 0.91 |
| Asian | 12 (2.56%) | 24 (2.41%) | 452 (3.88%) | 0.80 |
| Native Hawaiian or Pacific Islander | 3 (0.64%) | 1 (0.10%) | 42 (0.36%) | 0.07 |
| American Indian or Alaska Native | 14 (2.99%) | 11 (1.10%) | 207 (1.78%) | 0.009 |
| Other/Don't know | 25 (5.34%) | 32 (3.21%) | 642 (5.51%) | 0.05 |
| Hispanic Ethnicity | 55 (11.60%) | 75 (7.48%) | 1378 (11.72%) | 0.009 |
| MacArthur SES, Mean (SD) | 6.64 (1.54) | 7.08 (1.46) | 6.54 (1.68) | 0.0000 |
| Highest Educational Level |  |  |  |  |
| No high school degree | 0 | 3 (0.30%) | 79 (0.67%) | 0.0000 |
| High school graduate (or equivalent) | 16 (3.38%) | 15 (1.50%) | 622 (5.29%) |  |
| College degree (including associate's) | 279 (58.86%) | 486 (48.60%) | 6697 (56.97%) |  |
| Graduate degree | 171 (36.08%) | 488 (48.80%) | 4188 (35.62%) |  |
| Other | 8 (1.69%) | 8 (0.80%) | 168 (1.43%) |  |
| US resident | 461 (96.85%) | 984 (98.01%) | 11552 (97.72%) | 0.17 |
| Primary Employment |  |  |  |  |
| Healthcare | 95 (19.96%) | 187 (18.63%) | 2743 (23.20%) | 0.06 |
| Education | 71 (14.92%) | 154 (15.34%) | 1410 (11.93%) |  |
| Retail | 11 (2.31%) | 14 (1.39%) | 333 (2.82%) |  |
| Transportation | 12 (2.52%) | 12 (1.20%) | 213 (1.80%) |  |
| Arts, entertainment, and recreation | 13 (2.73%) | 33 (3.29%) | 298 (2.52%) |  |
| Hospitality and food services | 4 (0.84%) | 23 (2.29%) | 288 (2.44%) |  |
| Finance and insurance | 26 (5.46%) | 46 (4.58%) | 700 (5.92%) |  |
| Scientific and technical services | 30 (6.30%) | 100 (9.96%) | 792 (6.70%) |  |
| Utilities | 3 (0.63%) | 6 (0.60%) | 85 (0.72%) |  |
| Construction | 9 (1.89%) | 8 (0.80%) | 231 (1.95%) |  |
| Manufacturing | 14 (2.94%) | 23 (2.29%) | 324 (2.74%) |  |
| Other | 188 (39.50%) | 398 (39.64%) | 4405 (37.26%) |  |
| COVID-19 Variant wave |  |  |  |  |
| Initial | 236 (49.58%) | 244 (24.30%) | 5861 (49.56%) | <0.0001 |
| Alpha | 21 (4.41%) | 48 (4.78%) | 710 (6.00%) |  |
| Delta | 103 (21.64%) | 207 (20.62%) | 2202 (18.62%) |  |
| Omicron | 116 (24.37%) | 505 (50.30%) | 3052 (25.81%) |  |
| Days since COVID-19, median (IQR) | 360.0 (129, 506) | 129.0 (108, 343) |  | <0.0001 |
| Hospitalized due to COVID | 7 (1.47%) |  | 29 (0.25%) | 0.0001 |
| Max number of acute COVID-19 symptoms, mean (SD) | 5.44 ± 2.82 | 3.99 ± 2.39 | 4.08 ± 2.73 | <0.0001 |
| 1st Vaccine dose before COVID-19 | 213 (44.75%) | 710 (70.72%) | 5111 (43.22%) | <0.0001 |
| 1st Vaccine dose after COVID-19 | 216 (45.38%) | 211 (21.02%) | 3276 (27.70%) | <0.0001 |
| No reported vaccine | 47 (9.87%) | 83 (8.27%) | 3438 (29.07%) | 0.31 |
| Average days/week physical activity pre-COVID, mean (SD) | 2.49 ± 1.75 | 2.70 ± 1.90 | 2.52 ± 1.87 | 0.18 |
| Average days/week physical activity post-COVID (all), mean (SD) | 2.20 ± 1.73 | 2.62 ± 1.84 | 2.19 ± 1.84 | 0.0000 |
| Average days/week physical activity post-COVID (only those with pre-COVID data), mean (SD) | 2.14 ± 1.73 | 2.55 ± 1.83 | 2.17 ± 1.82 | 0.009 |
| Average hours of sleep/night pre-COVID, mean (SD) | 6.60 ± 0.93 | 6.85 ± 0.81 | 6.79 ± 0.92 | 0.0001 |
| Average hours of sleep/night post-COVID, mean (SD) | 6.62 ± 0.99 | 6.88 ± 0.86 | 6.83 ± 1.05 | 0.0000 |
| Average hours of sleep/night post-COVID (only those with pre-COVID data), mean (SD) | 6.68 ± 1.01 | 6.89 ± 0.85 | 6.89 ± 1.03 | 0.002 |
| BMI, mean (SD) | 30.58 (7.81) | 28.12 (6.91) | 29.47 (7.74) | <0.0001 |
| Tobacco use | 29 (6.09%) | 52 (5.18%) | 974 (8.75%) | 0.47 |
| Marijuana use | 30 (6.30%) | 70 (6.97%) | 953 (8.56%) | 0.63 |
| Alcoholic drinks/week pre-COVID, mean (SD) | 4.19 ± 5.71 | 4.81 ± 5.59 | 4.35 ± 5.65 | 0.15 |
| Alcoholic drinks/week post-COVID (only those with pre-COVID data), mean (SD) | 3.77 ± 5.28 | 4.41 ± 5.36 | 3.88 ± 5.35 | 0.12 |
| Alcoholic drinks/week post-COVID, mean (SD) | 2.99 ± 4.65 | 4.24 ± 5.26 | 3.28 ± 5.03 | <0.0001 |
| Financial insecurity pre-COVID | 66 (30.00%) | 92 (13.31%) | 806 (18.58%) | <0.0001 |
| Financial insecurity post-COVID (only those with pre-COVID data) | 60 (28.44%) | 72 (11.06%) | 577 (16.09%) | <0.0001 |
| Financial insecurity post-COVID | 149 (32.68%) | 131 (13.72%) | 1382 (20.64%) | <0.0001 |
| Anxiety (GAD-7) pre-COVID, mean (SD) | 5.83 ± 4.69 | 3.51 ± 3.50 | 4.40 ± 4.41 | <0.0001 |
| Anxiety (GAD-7) post-COVID (only those with pre-COVID data), mean (SD) | 5.41 ± 4.82 | 3.19 ± 3.57 | 3.94 ± 4.34 | <0.0001 |
| Anxiety (GAD-7) post-COVID, mean (SD) | 5.04 ± 4.66 | 3.08 ± 3.60 | 4.18 ± 4.53 | <0.0001 |
| Depression (PHQ-9) pre-COVID, mean (SD) | 6.27 ± 4.72 | 3.61 ± 3.56 | 4.56 ± 4.47 | <0.0001 |
| Depression (PHQ-9) post-COVID (only those with pre-COVID data), mean (SD) | 6.36 ± 5.13 | 3.50 ± 3.79 | 4.43 ± 4.58 | <0.0001 |
| Depression (PHQ-9) post-COVID, mean (SD) | 5.85 ± 4.91 | 3.31 ± 3.74 | 4.71 ± 4.85 | <0.0001 |
| Hypertension | 146 (30.67%) | 270 (26.89%) | 2954 (26.01%) | 0.11 |
| Diabetes | 40 (8.40%) | 68 (6.77%) | 850 (7.48%) | 0.23 |
| Coronary Artery Disease | 28 (5.88%) | 32 (3.19%) | 452 (3.98%) | 0.05 |
| Heart Failure | 6 (1.26%) | 10 (1.00%) | 126 (1.11%) | 0.75 |
| Stroke or TIA | 14 (2.94%) | 20 (1.99%) | 236 (2.08%) | 0.002 |
| Atrial Fibrillation | 29 (6.09%) | 41 (4.08%) | 424 (3.73%) | 0.06 |
| Obstructive Sleep apnea | 85 (17.86%) | 124 (12.35%) | 1474 (12.98%) | 0.005 |
| Chronic Obstructive Pulmonary Disease | 27 (5.67%) | 15 (1.49%) | 275 (2.42%) | <0.0001 |
| Asthma | 68 (14.29%) | 85 (8.47%) | 1237 (10.89%) | 0.0009 |
| Cancer | 28 (5.88%) | 74 (7.37%) | 544 (4.79%) | 0.48 |
| Immunodeficiency | 29 (6.09%) | 20 (1.99%) | 396 (3.49%) | 0.0002 |
| HIV | 4 (0.84%) | 5 (0.50%) | 71 (0.63%) | 0.13 |
| Pregnant | 3 (0.63%) | 8 (0.80%) | 152 (1.34%) | 0.90 |
| COVID prior to baseline survey | 237 (49.79%) | 274 (27.29%) | 6432 (54.39%) | <0.0001 |

The most common Long COVID symptom was fatigue, reported by 230/476 (48.3%) (Figure 1). Other common symptoms included shortness of breath (109, 22.9%), confusion (108, 22.7%) headache (103, 21.6%), and altered taste or smell (98, 20.6%). A minority of participants (62/476, 13.0%) reported at least one severe or very severe symptom. Health care contact about Long COVID was reported by 219/476 (46.0%) of participants. Missing work or school due to Long COVID was reported by 124 (26.1%) participants, with 57 (12.0%) missing 1-5 days, 24 (5.0%) missing 6-10 days, and 43 (9.0%) missing 11 days for more.

**Figure 1.**
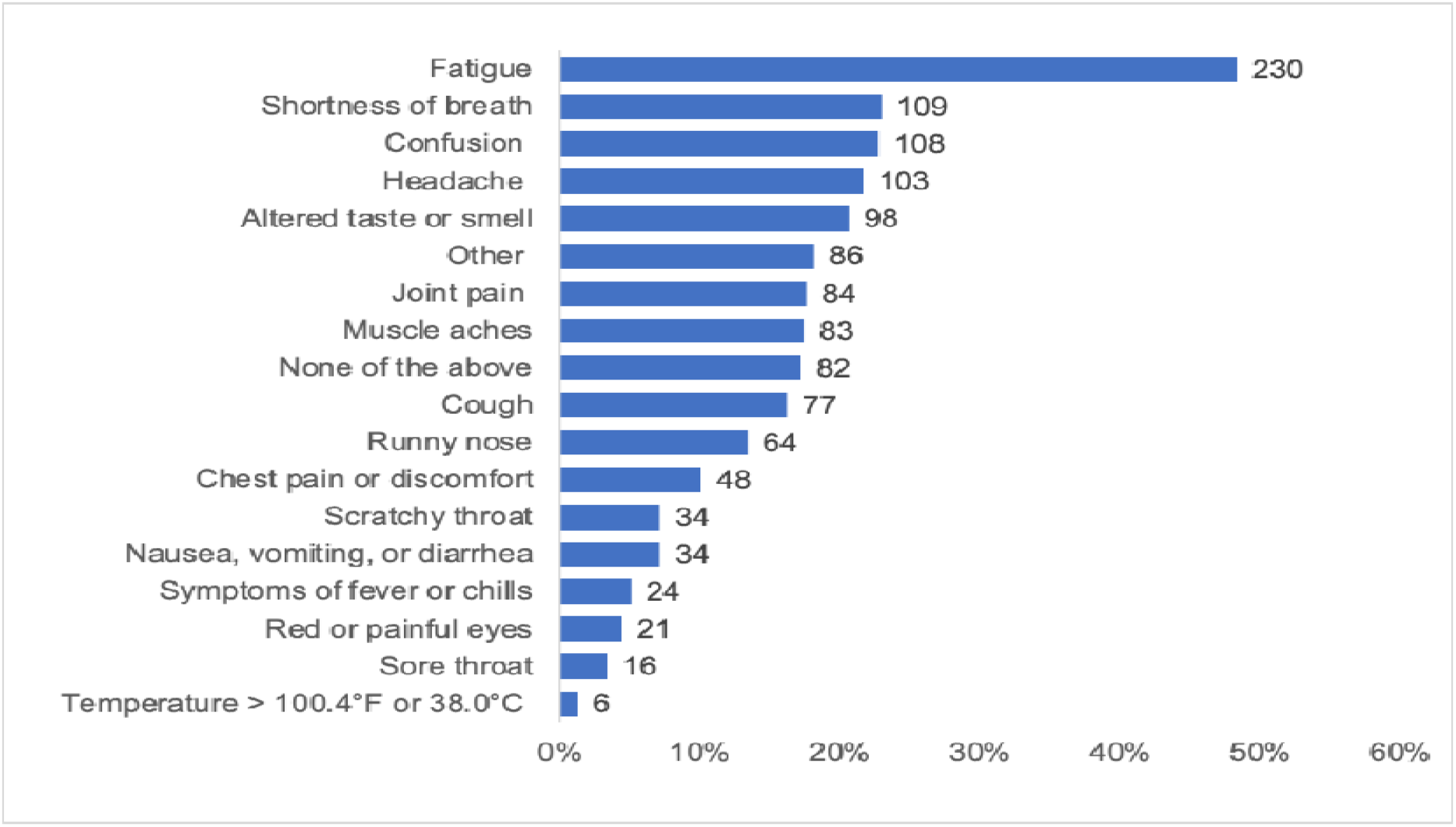
Patient-reported symptoms of Long COVID among people reporting symptoms at least 1 month after COVID-19 (N = 476). Numbers to the right of bars represent number of participants reporting that symptom. Participants could report more than one symptom. Number and proportion with each symptom.

**Figure 2.**
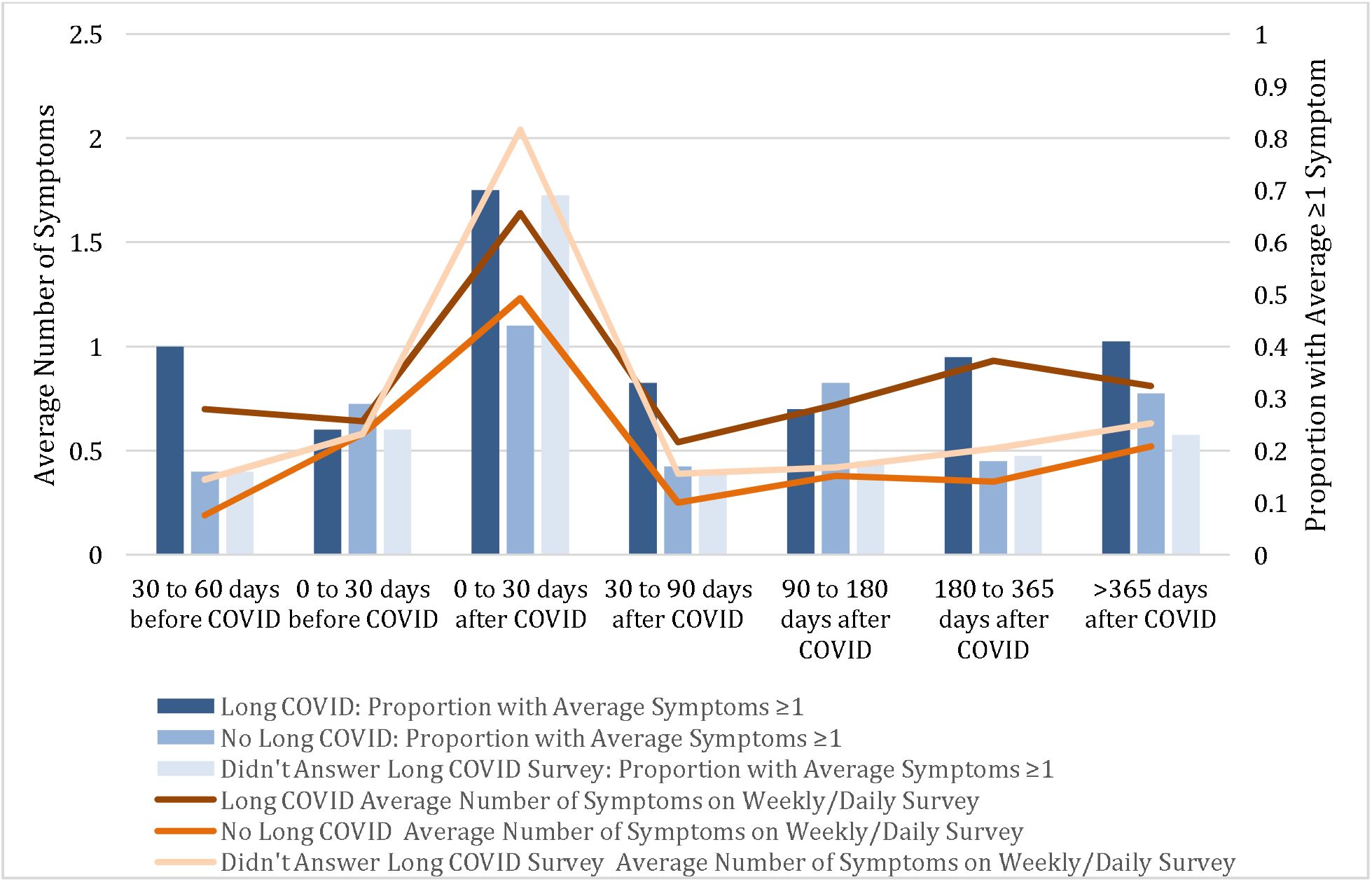
Average Number of Symptoms and Proportion with Symptoms on the Weekly/Daily Surveys During Each Time Period. We found higher proportions with symptoms and a higher number of symptoms reported among those with Long COVID. Of note, symptoms queried on the daily/weekly surveys included whether participants had one or more of the following: a scratchy throat, a painful sore throat, a cough, a runny nose, symptoms of fever or chills, a temperature >100.4°F or 38.0°C, muscle aches, nausea, vomiting, or diarrhea, shortness of breath, unable to taste or smell, and red or painful eyes. These symptoms are more typical during acute infection so some individuals with Long COVID did not have any of these symptoms but still reported Long COVID symptoms.

Long COVID symptoms lasted for varying durations, with nearly half of participants reporting ongoing symptoms (227/476, 47.7%) (Table 2). Of participants with COVID-19 at least 1 year prior to survey completion who reported experiencing Long COVID symptoms, 133/237 (56.1%) reported still experiencing symptoms at the time of survey completion.

**Table 2.**
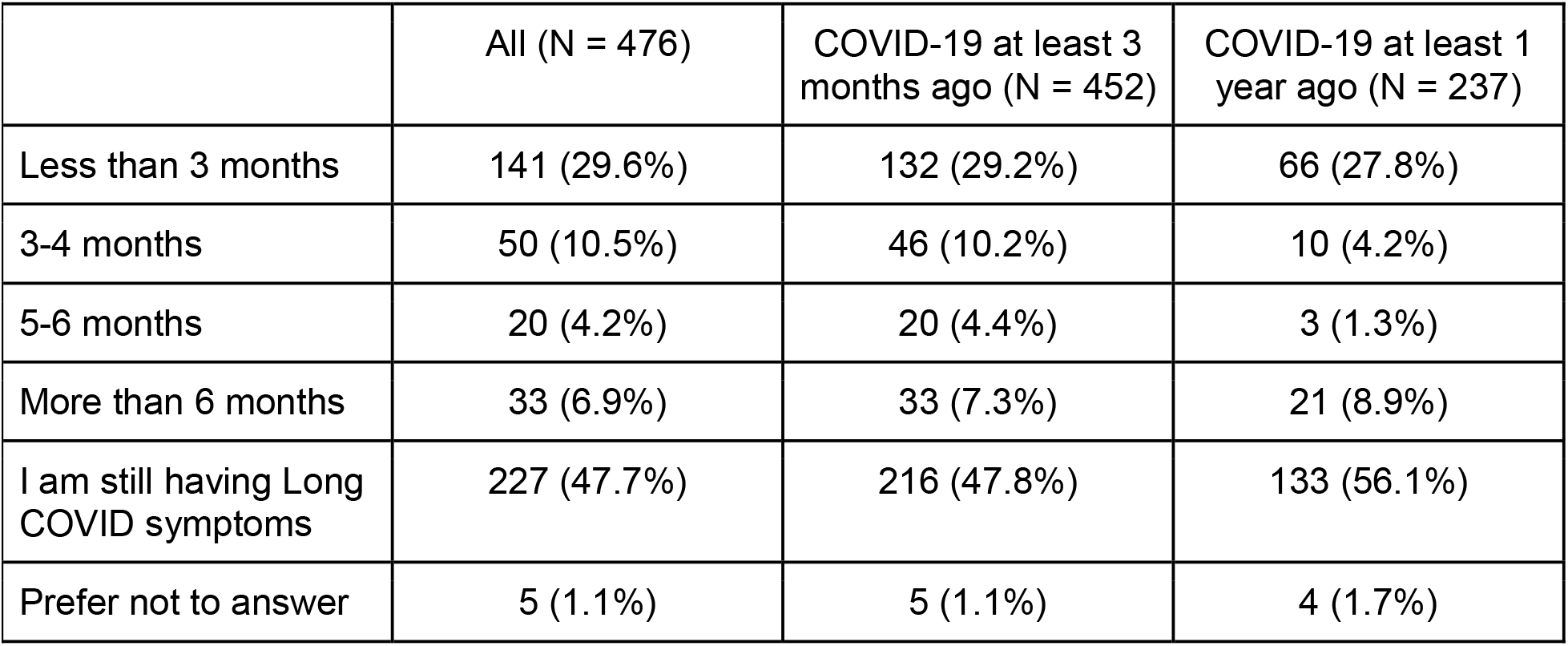
Duration of Long COVID symptoms among people reporting symptoms at least 1 month after COVID-19. Number and proportion reporting each duration of symptoms and those reporting ongoing symptoms by time of initial infection (any time, more than 3 months prior to survey, and more than 1 year prior to survey). Note - Having symptoms longer than 3 months is most consistent with the WHO definition.^14^

In all three multivariable models, the number of acute COVID-19 symptoms during initial infection was associated with prevalent Long COVID symptoms with 1.3 times higher odds per additional acute symptom (95%CI 1.20-1.40 for Model 3; Table 3). Variant wave was also associated with prevalent Long COVID symptoms, with later wave participants less likely to have Long COVID symptoms despite shorter follow-up time (median follow up 360 days [IQR 129, 506] among symptomatic, 129 days [IQR 108, 343] among asymptomatic). When models were further adjusted for vaccination before or after COVID, ethnicity and social determinants of health (subjective socioeconomic status, highest level of education, employment in healthcare), Hispanic ethnicity and lower subjective socioeconomic status were associated with Long COVID, with odds ratios of 1.73 (95%CI 1.02-2.83) and 0.81 per unit higher (95%CI 0.73 to 0.91), respectively (Model 2; Table 3). Vaccination status was not statistically significantly associated with Long COVID symptoms (OR 0.81 for pre-infection, 95%CI 0.44-1.49; 1.57 for post-infection vaccination 95%CI 0.60-4.13). After additional adjustment for anxiety, depression, and financial insecurity, pre-existing depression (OR 1.08 per point on PHQ-8, 95%CI 1.01-2.16) and financial insecurity (OR 1.64, 95%CI 1.02-2.63) were associated with Long COVID.

**Table 3.** Multivariable models of factors potentially associated with Long COVID. **Model 1** includes age, COVID-19 wave, number of initial symptoms, sex, myocardial infarction, stroke, atrial fibrillation, sleep apnea, COPD or asthma, immunodeficiency. **Model 2** includes Model 1 factors, plus receipt of COVID-19 vaccination before or after COVID-19 diagnosis, ethnicity, subjective socioeconomic status, highest level of education, and primary employment. **Model 3** includes Model 2 factors, plus pre-COVID-19 depressive symptoms, anxiety symptoms, and financial insecurity.

|  | Model 1<br>N = 1024 |  | Model 2<br>N = 1021 |  | Model 3<br>N = 905 |  |
| --- | --- | --- | --- | --- | --- | --- |
| Variable | OR<br>(95%CI) | p-value | OR<br>(95%CI) | p-value | OR<br>(95%CI) | p-value |
| Age | 1.00 (0.99, 1.01) | 0.90 | 1.00 (0.99, 1.02) | 0.66 | 1.01 (0.99, 1.03) | 0.14 |
| Wave |  |  |  |  |  |  |
| Initial | Reference | Reference | Reference | Reference | Reference | Reference |
| Alpha | 0.44 (0.17, 1.10) | 0.08 | 0.64 (0.23, 1.77) | 0.39 | 1.20 (0.37, 3.90) | 0.76 |
| Delta | 0.33 (0.21, 0.53) | <.001 | 0.48 (0.22, 1.06) | 0.07 | 0.57 (0.21, 1.50) | 0.25 |
| Omicron | 0.17 (0.11, 0.26) | <.001 | 0.25 (0.12, 0.54) | <.001 | 0.37 (0.15, 0.90) | 0.04 |
| Number of initial symptoms (per symptom) | 1.27 (1.20, 1.35) | <.001 | 1.27 (1.20, 1.35) | <.001 | 1.30 (1.20, 1.40) | <.001 |
| Female sex | 1.24 (0.88, 1.77) | 0.22 | 1.16 (0.80, 1.66) | 0.44 | 0.86 (0.57, 1.29) | 0.47 |
| Myocardial Infarction | 0.58 (0.27, 1.25) | 0.16 | 0.63 (0.29, 1.37) | 0.24 | 0.57 (0.23, 1.39) | 0.21 |
| Stroke | 0.82 (0.30, 2.19) | 0.69 | 0.96 (0.34, 2.68) | 0.94 | 1.46 (0.47, 4.52) | 0.51 |
| Atrial Fibrillation | 0.51 (0.26, 1.01) | 0.05 | 0.47 (0.23, 0.96) | 0.04 | 0.61 (0.28, 1.31) | 0.20 |
| Sleep apnea | 1.00 (0.63, 1.58) | 0.99 | 1.07 (0.67, 1.70) | 0.79 | 1.14 (0.68, 1.89) | 0.63 |
| COPD or Asthma | 0.79 (0.17, 3.68) | 0.76 | 0.87 (0.18, 4.14) | 0.86 | 0.89 (0.17, 4.80) | 0.89 |
| Immunodeficiency | 0.54 (0.27, 1.07) | 0.08 | 0.51 (0.25, 1.03) | 0.06 | 0.57 (0.26, 1.28) | 0.18 |
| Vaccination before COVID-19 | N/A | N/A | 0.85 (0.49, 1.49) | 0.57 | 0.81 (0.44, 1.49) | 0.50 |
| Vaccination after COVID-19 | N/A | N/A | 1.41 (0.65, 3.06) | 0.39 | 1.57 (0.60, 4.13) | 0.36 |
| Hispanic ethnicity | N/A | N/A | 1.70 (1.02, 2.83) | 0.04 | 1.73 (0.95, 3.14) | 0.07 |
| Subjective socioeconomic status (per unit higher) | N/A | N/A | 0.81 (0.73, 0.91) | <.001 | 0.90 (0.79, 1.03) | 0.12 |
| Highest education | N/A | N/A |  |  |  |  |
| No high school | N/A | N/A | N/A | N/A | N/A | N/A |
| High school graduate | N/A | N/A | 0.72 (0.12, 4.40) | 0.72 | 0.69 (0.08, 6.03) | 0.74 |
| College degree | N/A | N/A | 0.84 (0.22, 3.25) | 0.80 | 1.17 (0.23, 5.97) | 0.85 |
| Graduate degree | N/A | N/A | 0.73 (0.19, 2.86) | 0.65 | 0.99 (0.19, 5.12) | 0.99 |
| Other | N/A | N/A | Reference | Reference | Reference | Reference |
| Healthcare worker | N/A | N/A | 1.02 (0.98, 1.06) | 0.28 | 1.02 (0.98, 1.07) | 0.25 |
| Pre-COVID-19 Depressive Symptoms | N/A | N/A | N/A | N/A | 1.08 (1.01, 1.16) | 0.03 |
| Pre-COVID-19 Anxiety Symptoms | N/A | N/A | N/A | N/A | 1.04 (0.97, 1.12) | 0.25 |
| Pre-COVID-19 financial insecurity | N/A | N/A | N/A | N/A | 1.64 (1.02, 2.63) | 0.04 |

Among those with pre-COVID baseline data, physical activity, alcohol intake, and anxiety were lower after COVID regardless of Long COVID status (eTable 1). There was a greater decrease in frequency of physical activity after COVID-19 among those with Long COVID (difference in change between symptomatic and asymptomatic: 0.19 days/week, 95%CI 0.04-0.35, p=0.02), but no differences in change in sleep duration, alcohol intake, anxiety, or depression (eTable 1).

Among those who responded to daily or weekly survey requests including non-respondents to the cross-sectional Long COVID survey, we plotted the average number of symptoms reported (line) and the proportion who reported an average of one or more symptom over all surveys answered (bars) during time periods before and after SARS-CoV-2 infection for those infected (Figure 4). The estimated proportion reporting an average of one or more symptom on all surveys was highest in the 0-30 days after acute infection in all groups. The estimated proportion with an average of 1 or more symptom across completed surveys for each time period was generally higher among those reporting Long COVID symptoms (at 180-365 days: 38% vs 18% without Long COVID symptoms) and substantially higher than among SARS-CoV-2 uninfected individuals (n=57,415, 11%). Similarly, the average number of symptoms was higher among those reporting Long COVID (at 180-365 days: 0.81±1.81) compared to those not reporting Long COVID symptoms (0.35±0.98) and those without SARS-CoV-2 infection (n=57,415, 0.32±0.60).

### Sensitivity Analyses

First, we considered whether selection bias bias from those who joined the study and completed baseline surveys after SARS-CoV-2 infection impacted the findings. Among 969 individuals with pre-infection baseline surveys, 239 (25%) reported Long COVID symptoms compared to 237/511 who completed baseline surveys after SARS-CoV-2 infection (46%; p<0.0001), which suggests that individuals with Long COVID symptoms after SARS-CoV-2 infection were more likely to participate than those without Long COVID symptoms, consistent with selection bias. Importantly, mean levels of reported physical activity, sleep duration, alcohol intake, financial insecurity, depression, and anxiety were similar between those with baseline surveys obtained prior to SARS-CoV-2 infection and those who joined the cohort after infection, suggestive that differential recall is unlikely to bias these variables (eTable 2). The general pattern of results for the multivariable models were unchanged when only individuals included with baseline surveys prior to infection (n=239 with and = 730 without symptoms) were included (eTable 3).

Second, we considered only persistent symptoms (n=227) compared to those no symptoms (n=1004), excluding those whose symptoms resolved (eTable 3). Overall results were similar with one exception: female sex was associated with persistent symptoms (OR 1.34, 95%CI 1.21-1.49; eTable 3). We also compared those with severe or very severe symptoms (n=62) to those without symptoms (n=1004), with no substantive differences in our findings (eTable 3).

## Discussion

In this cross-sectional assessment of mostly non-hospitalized individuals who reported prior SARS-CoV-2 infection in the COVID Citizen Science online cohort, persistent symptoms including fatigue, shortness of breath, headache, brain fog/confusion, and altered taste/smell were highly prevalent. A minority of participants reported severe or very severe symptoms, but half of participants infected more than 1 year earlier reported ongoing symptoms. We found that pre-infection socioeconomic status, financial insecurity, and depression assessed prior to SARS-CoV-2 infection were associated with Long COVID symptoms. We also found that the number of symptoms during acute infection was associated with reporting Long COVID symptoms independent of vaccination and variant wave, and that more recent variant waves are associated with lower odds of Long COVID even after adjusting for vaccine status.

### Comparison of Symptom Patterns and Persistence

Our findings of common symptoms (fatigue, shortness of breath, confusion, and headache) are consistent with prior reports.^1-3^ Similarly, we found that symptoms were persistent for more than 12 months among approximately half of those infected with SARS-CoV-2 who reported symptoms lasting at least one month. This is consistent with the prior literature that symptoms present for more than 3 months tend not to self-resolve,^1^ but higher than estimates that only 15% of individuals with symptoms at 3 months continue to have symptoms beyond 1 year.^2^

### Risk of Long COVID by Symptoms and Variant Wave

The number of symptoms during acute infection was associated with Long COVID consistent with prior reports that acute illness severity is associated with Long COVID,^8^ and raises the question of whether reducing acute symptoms through acute treatment might modify the risk of developing Long COVID. A second interesting finding is that the variant wave is associated with Long COVID symptoms even with adjustment for timing of vaccination (pre-infection, post-infection, or not vaccinated) and number of symptoms during acute infection; more recent variants were associated with lower odds of Long COVID. One prior study suggested that there may be some subtle differences in Long COVID symptoms by variant wave (more dyspnea with ancestral strain, more neuropsychiatric and myalgic symptoms with Alpha, and hair loss with Delta for example).^23,24^ Our findings are consistent with three prior studies that suggested that there may be a lower prevalence of Long COVID with the more recent variants (Epsilon, Omicron).^15,25,26^

### Demographics, Social Determinants of Health and Long COVID

Even within a relatively homogenous cohort, we found that Hispanic ethnicity, lower socioeconomic status and financial insecurity were associated with Long COVID symptoms. In contrast to prior reports, we did not identify female sex as associated with Long COVID symptoms after adjustment, except in sensitivity analysis of persistent symptoms.^10,28^ Despite extensive research documenting the role of adverse social determinants of health increasing risk of acute SARS-CoV-2 infection, there are limited prior data regarding associations between social determinants and Long COVID. Consistent with our study, one prior study found that financial concerns were associated with worse health utility and quality of life among those recovering from COVID-19.^27^ Similarly, an online survey-based study found that graduate education and urban residence were associated with lower odds of Long COVID.^15^ The implications are that clinical trials of potential therapeutics should make intentional efforts to include those at highest risk, including those of lower socioeconomic status, and social determinants of health should be considered in public health approaches to address Long COVID.

### Depression, Anxiety and Long COVID

In our adjusted analysis, pre-COVID depression was associated with Long COVID symptoms. Importantly, we found that anxiety and depression scores did not increase after COVID among those with pre-infection baseline surveys, suggesting that individuals with depression may be at elevated risk of Long COVID. One prior study from three large cohorts identified that depression, anxiety, perceived stress, and loneliness measured before the pandemic were associated with post-COVID conditions, although we found that anxiety decreased among those with Long COVID.^29^ Our findings are consistent with studies that lack pre-infection assessments that have found concurrent depression or anxiety to be associated with Long COVID symptoms.^10,11,28,30,31^ Further research into mechanisms of how depression may be an antecedent factor to Long COVID are needed. Fluvoxamine, a selective serotonin reuptake inhibitor (SSRI), may have protective effects in acute SARS-CoV-2 infection,^32,33^ but understanding whether treatment with antidepressants or naltrexone^34^ may prevent or treat Long COVID requires clinical trials.^35^

### Strengths and Limitations

Strengths of this online cohort are a large sample size, data collection prior to infection in many participants, and data from participants infected during different variant waves; most prior studies included predominantly individuals infected with earlier variants. The primary limitation arises from responder bias: those with Long COVID are more likely to respond to surveys although over 1000 individuals without symptoms also responded. This may induce bias in our estimate of Long COVID symptom prevalence but is less likely to bias ascertainment of factors associated with Long COVID. The second limitation is that a subset of individuals joined the study after SARS-CoV-2 infection, limiting the ability for prospective comparisons to those who had already joined the study. Symptoms may be misclassified, and some may be attributable to specific medical conditions rather than Long COVID. We did not assess the effect of repeat SARS-CoV-2 infection. Finally, external generalizability may be limited as the study sample overrepresented those who identify as white, female, and of higher socioeconomic status.

## Conclusions

In conclusion, in this cross-sectional assessment of Long COVID symptoms within an online cohort, we found that Long COVID symptoms were highly prevalent and commonly persisted, consistent with prior reports. We found that having more symptoms during acute infection, lower socioeconomic status, financial stress, and pre-COVID depression were associated with Long COVID. Finally, Long COVID symptoms were less prevalent among those infected with recent variants even accounting for vaccination status and acute symptoms.

## Supporting information

STROBE Checklist

## Data Availability

Data produced in this study are available on reasonable request to the COVID Citizen Science leadership committee.

## Author Disclosures

Dr. Peluso has received consulting fees from Gilead Sciences and AstraZeneca, outside the submitted work. Dr. Beatty was formerly employed by Apple Inc. (2018-2019), held stock in Apple Inc. (2019-2021), and receives consulting fees from AHRQ-funded projects unrelated to this work. Dr. Djibo is employed by CVS Health. Other authors have not disclosed any conflicts of interest.

## Funding

Eureka Research Platform is supported by NIH/NIBIB 3U2CEB021881-05S1. The COVID-19 Citizen Science Study is supported by Patient-Centered Outcomes Research Institute (PCORI) contract COVID-2020C2-10761 and Bill and Melinda Gates Foundation contract INV-017206. Dr. Durstenfeld is supported by NIH/NHLBI grant K12HL143961. Dr. Peluso is supported by NIH/NIAIN grant K23AI157875.

## Notes

### Author Declarations

University of California, San Francisco Institutional Review Board reviewed this study and gave ethical approval (#17-21879).

## References

1. Peluso MJ, Kelly JD, Lu S, et al. Persistence, Magnitude, and Patterns of Postacute Symptoms and Quality of Life Following Onset of SARS-CoV-2 Infection: Cohort Description and Approaches for Measurement. Open Forum Infect Dis. 2022;9(2):ofab640.

2. Collaborators GBoDLC. Estimated Global Proportions of Individuals With Persistent Fatigue, Cognitive, and Respiratory Symptom Clusters Following Symptomatic COVID-19 in 2020 and 2021. JAMA. 2022.

3. Groff D, Sun A, Ssentongo AE, et al. Short-term and Long-term Rates of Postacute Sequelae of SARS-CoV-2 Infection: A Systematic Review. JAMA Netw Open. 2021;4(10):e2128568.

4. Hirschtick JL, Titus AR, Slocum E, et al. Population-Based Estimates of Post-acute Sequelae of Severe Acute Respiratory Syndrome Coronavirus 2 (SARS-CoV-2) Infection (PASC) Prevalence and Characteristics. Clin Infect Dis. 2021;73(11):2055–2064.

5. Taquet M, Dercon Q, Luciano S, Geddes JR, Husain M, Harrison PJ. Incidence, co-occurrence, and evolution of long-COVID features: A 6-month retrospective cohort study of 273,618 survivors of COVID-19. PLoS Med. 2021;18(9):e1003773.

6. Piotr DA, Gaughan PC. Technical article: Updated estimates of the prevalence of post-acute symptoms among people with coronavirus (COVID-19) in the UK: 26 April 2020 to 1 August 2021. In: Statistics UOfN, ed. London 2021.

7. Yomogida K, Zhu S, Rubino F, Figueroa W, Balanji N, Holman E. Post-Acute Sequelae of SARS-CoV-2 Infection Among Adults Aged >/=18 Years -Long Beach, California, April 1-December 10, 2020. MMWR Morb Mortal Wkly Rep. 2021;70(37):1274–1277.

8. Xie Y, Bowe B, Al-Aly Z. Burdens of post-acute sequelae of COVID-19 by severity of acute infection, demographics and health status. Nat Commun. 2021;12(1):6571.

9. Al-Aly Z, Bowe B, Xie Y. Long COVID after breakthrough SARS-CoV-2 infection. Nature Medicine. 2022.

10. Hastie CE, Lowe DJ, McAuley A, et al. Outcomes among confirmed cases and a matched comparison group in the Long-COVID in Scotland study. Nat Commun. 2022;13(1):5663.

11. Li D, Liao X, Liu Z, et al. Healthy Outcomes of Patients with COVID-19 Two Years after the Infection: A Prospective Cohort Study. Emerg Microbes Infect. 2022:1–32.

12. Huang Y, Pinto MD, Borelli JL, et al. COVID Symptoms, Symptom Clusters, and Predictors for Becoming a Long-Hauler Looking for Clarity in the Haze of the Pandemic. Clin Nurs Res. 2022;31(8):1390–1398.

13. Hill E, Mehta H, Sharma S, et al. Risk Factors Associated with Post-Acute Sequelae of SARS-CoV-2 in an EHR Cohort: A National COVID Cohort Collaborative (N3C) Analysis as part of the NIH RECOVER program. medRxiv. 2022.

14. Al-Aly Z, Xie Y, Bowe B. High-dimensional characterization of post-acute sequelae of COVID-19. Nature. 2021;594(7862):259–264.

15. Perlis RH, Santillana M, Ognyanova K, et al. Prevalence and Correlates of Long COVID Symptoms Among US Adults. JAMA Netw Open. 2022;5(10):e2238804.

16. Wu Q, Ailshire JA, Crimmins EM. Long COVID and symptom trajectory in a representative sample of Americans in the first year of the pandemic. Sci Rep. 2022;12(1):11647.

17. Beatty AL, Peyser ND, Butcher XE, et al. The COVID-19 Citizen Science Study: Protocol for a Longitudinal Digital Health Cohort Study. JMIR Res Protoc. 2021;10(8):e28169.

18. von Elm E, Altman DG, Egger M, Pocock SJ, Gøtzsche PC, Vandenbroucke JP. The Strengthening the Reporting of Observational Studies in Epidemiology (STROBE) statement: guidelines for reporting observational studies. Ann Intern Med. 2007;147(8):573–577.

19. Lauring AS, Tenforde MW, Chappell JD, et al. Clinical severity of, and effectiveness of mRNA vaccines against, covid-19 from omicron, delta, and alpha SARS-CoV-2 variants in the United States: prospective observational study. BMJ. 2022;376:e069761.

20. Kroenke K, Strine TW, Spitzer RL, Williams JB, Berry JT, Mokdad AH. The PHQ-8 as a measure of current depression in the general population. J Affect Disord. 2009;114(1-3):163–173.

21. Spitzer RL, Kroenke K, Williams JBW, Löwe B. A Brief Measure for Assessing Generalized Anxiety Disorder. Archives of Internal Medicine. 2006;166(10).

22. Adler N, Singh-Manoux A, Schwartz J, Stewart J, Matthews K, Marmot MG. Social status and health: a comparison of British civil servants in Whitehall-II with European- and African-Americans in CARDIA. Soc Sci Med. 2008;66(5):1034–1045.

23. Fernández-de-Las-Peñas C, Cancela-Cilleruelo I, Rodríguez-Jiménez J, et al. Associated-Onset Symptoms and Post-COVID-19 Symptoms in Hospitalized COVID-19 Survivors Infected with Wuhan, Alpha or Delta SARS-CoV-2 Variant. Pathogens. 2022;11(7).

24. Spinicci M, Graziani L, Tilli M, et al. Infection with SARS-CoV-2 Variants Is Associated with Different Long COVID Phenotypes. Viruses. 2022;14(11).

25. Antonelli M, Pujol JC, Spector TD, Ourselin S, Steves CJ. Risk of long COVID associated with delta versus omicron variants of SARS-CoV-2. Lancet. 2022;399(10343):2263–2264.

26. Morioka S, Tsuzuki S, Suzuki M, et al. Post COVID-19 condition of the Omicron variant of SARS-CoV-2. J Infect Chemother. 2022;28(11):1546–1551.

27. Case KR, Wang CP, Hosek MG, et al. Health-related quality of life and social determinants of health following COVID-19 infection in a predominantly Latino population. J Patient Rep Outcomes. 2022;6(1):72.

28. Sneller MC, Liang CJ, Marques AR, et al. A Longitudinal Study of COVID-19 Sequelae and Immunity: Baseline Findings. Ann Intern Med. 2022;175(7):969–979.

29. Wang S, Quan L, Chavarro JE, et al. Associations of Depression, Anxiety, Worry, Perceived Stress, and Loneliness Prior to Infection With Risk of Post-COVID-19 Conditions. JAMA Psychiatry. 2022.

30. Mazza MG, Palladini M, Villa G, De Lorenzo R, Rovere Querini P, Benedetti F. Prevalence, trajectory over time, and risk factor of post-COVID-19 fatigue. J Psychiatr Res. 2022;155:112–119.

31. Margalit I, Yelin D, Sagi M, et al. Risk factors and multidimensional assessment of long COVID fatigue: a nested case-control study. Clin Infect Dis. 2022.

32. Lenze EJ, Mattar C, Zorumski CF, et al. Fluvoxamine vs Placebo and Clinical Deterioration in Outpatients With Symptomatic COVID-19: A Randomized Clinical Trial. Jama. 2020;324(22):2292–2300.

33. Reis G, Dos Santos Moreira-Silva EA, Silva DCM, et al. Effect of early treatment with fluvoxamine on risk of emergency care and hospitalisation among patients with COVID-19: the TOGETHER randomised, platform clinical trial. Lancet Glob Health. 2022;10(1):e42–e51.

34. O’Kelly B, Vidal L, McHugh T, Woo J, Avramovic G, Lambert JS. Safety and efficacy of low dose naltrexone in a long covid cohort; an interventional pre-post study. Brain Behav Immun Health. 2022;24:100485.

35. Bonnet U, Juckel G. COVID-19 Outcomes: Does the Use of Psychotropic Drugs Make a Difference? Accumulating Evidence of a Beneficial Effect of Antidepressants-A Scoping Review. J Clin Psychopharmacol. 2022;42(3):284–292.

