## Supplemental Tables for "Factors Associated with Long Covid Symptoms in an Online Cohort Study"

**eTable 1. Change from Pre-COVID to Post-COVID among those Reporting LONG COVID Symptoms, Not Reporting Long COVID Symptoms, and Difference Between Groups**

|  | **Change from Pre-COVID to Post-COVID among those with Long COVID Symptoms** | | | **Change from Pre-COVID to Post-COVID among those without Long COVID Symptoms** | | | **Difference in Change between those with and without Long COVID** | |
| --- | --- | --- | --- | --- | --- | --- | --- | --- |
|  | N | Mean change (95%CI) | P value | N | Mean change (95%CI) | P value | Mean (95%CI) | P value |
| Average days/week physical activity | 169 | -0.35 (-0.50, -0.20) | <.0001 | 636 | -0.16 (-0.23, -0.09) | <.0001 | -0.19 (-0.35, -0.04) | 0.016 |
| Average hours sleep/night | 230 | 0.07 (0.001, 0.14) | 0.047 | 717 | 0.04 (0.01, 0.07) | 0.017 | 0.03 (-0.04, 0.11) | 0.39 |
| Alcoholic drinks/week | 223 | -0.40 (-0.79, -0.01) | 0.044 | 683 | -0.37 (-0.54, -0.20) | <.0001 | -0.03 (-0.46, 0.40) | 0.89 |
| Average Anxiety (GAD-7) | 212 | -0.50 (-0.85, -0.15) | 0.006 | 656 | -0.25 (-0.42, -0.09) | 0.003 | -0.25 (-0.64, 0.14) | 0.21 |
| Average Depression (PHQ-8) | 212 | 0.04 (-0.29, 0.38) | 0.80 | 656 | -0.07 (-0.22, 0.07) | 0.32 | 0.12 (-0.24, 0.47) | 0.53 |

eTable 1 Legend: Change in lifestyle factors, anxiety, and depression scores from pre-COVID to post-COVID among those with and without self-reported Long COVID symptoms on cross-sectional surveys and difference in change between those with and without Long COVID symptoms. P-values are for paired t-tests for within individual change in each group and unpaired t-test for the difference in change.

**eTable 2.** Pre- and Post- COVID lifestyle factors, Anxiety, and Depression Only Including those with Pre COVID baseline surveys

|  | Long COVID | | No Long COVID | | Non-Respondent | |  |
| --- | --- | --- | --- | --- | --- | --- | --- |
|  | N | Mean±SD | N | Mean±SD | N | Mean±SD | P value |
| Average days/week physical activity pre-COVID, Mean +/- SD | 169 | 2.49 ± 1.75 | 636 | 2.70 ± 1.90 | 3678 | 2.52 ± 1.87 | 0.1797 |
| Average days/week physical activity post-COVID (pre), Mean +/- SD | 169 | 2.14 ± 1.73 | 636 | 2.55 ± 1.83 | 3674 | 2.17 ± 1.82 | 0.0092 |
| Average days/week physical activity post-COVID, Mean +/- SD | 476 | 2.20 ± 1.73 | 1004 | 2.62 ± 1.84 | 7587 | 2.19 ± 1.84 | 0.0000 |
| Average sleep/week pre-COVID, Mean +/- SD | 230 | 6.60 ± 0.93 | 717 | 6.85 ± 0.81 | 4902 | 6.79 ± 0.92 | 0.0001 |
| Average sleep/week post-COVID (pre), Mean +/- SD | 230 | 6.68 ± 1.01 | 717 | 6.89 ± 0.85 | 4894 | 6.89 ± 1.03 | 0.0017 |
| Average sleep/week post-COVID, Mean +/- SD | 476 | 6.62 ± 0.99 | 1004 | 6.88 ± 0.86 | 9039 | 6.83 ± 1.05 | 0.0000 |
| Alcoholic drinks/week pre-COVID, Mean +/- SD | 232 | 4.19 ± 5.71 | 721 | 4.81 ± 5.59 | 5084 | 4.35 ± 5.65 | 0.1454 |
| Alcoholic drinks/week post-COVID (pre), Mean +/- SD | 223 | 3.77 ± 5.28 | 683 | 4.41 ± 5.36 | 4039 | 3.88 ± 5.35 | 0.1209 |
| Alcoholic drinks/week post-COVID, Mean +/- SD | 467 | 2.99 ± 4.65 | 966 | 4.24 ± 5.26 | 10739 | 3.28 ± 5.03 | 0.0000 |
| Average Anxiety (GAD-7) pre-COVID, mean +/- SD | 221 | 5.83 ± 4.69 | 692 | 3.51 ± 3.50 | 4378 | 4.40 ± 4.41 | 0.0000 |
| Average Anxiety (GAD-7) post-COVID (pre), mean +/- SD | 212 | 5.41 ± 4.82 | 656 | 3.19 ± 3.57 | 3648 | 3.94 ± 4.34 | 0.0000 |
| Average Anxiety (GAD-7) post-COVID, mean +/- SD | 456 | 5.04 ± 4.66 | 960 | 3.08 ± 3.60 | 6805 | 4.18 ± 4.53 | 0.0000 |
| Average Depression (PHQ-9) pre-COVID, mean +/- SD | 221 | 6.27 ± 4.72 | 692 | 3.61 ± 3.56 | 4375 | 4.56 ± 4.47 | 0.0000 |
| Average Depression (PHQ-9) post-COVID (pre), mean +/- SD | 212 | 6.36 ± 5.13 | 656 | 3.50 ± 3.79 | 3634 | 4.43 ± 4.58 | 0.0000 |
| Average Depression (PHQ-8) post-COVID, mean +/- SD | 456 | 5.85 ± 4.91 | 960 | 3.31 ± 3.74 | 6787 | 4.71 ± 4.85 | 0.0000 |

eTable 2 Table Legend: p-values reported are from t-tests to compare means between those with and without Long COVID.

**eTable 3.** Sensitivity Analyses of factors potentially associated with Long COVID.

|  | **Primary Analysis (Model 3): Any Long COVID symptom vs No Symptoms**  **(n=476 with, 1004 without)** | **Only with Pre-COVID Baseline Survey**  **(n=239 with, 730 without)** | **Persistent Symptoms vs No Symptoms**  **(n=227 with, 1004 without)** | **At Least 1 Severe/Very Severe Symptom vs No Symptoms**  **(n=62 with, 1004 without)** |
| --- | --- | --- | --- | --- |
| **Age, per year** | 1.01 (0.99, 1.03) | 1.01 (1.00, 1.03) | 1.02 (1.00-1.04) | 1.07 (1.01-1.13) |
| **Female sex** | 0.86 (0.57, 1.29) | 0.94 (0.61, 1.45) | 1.34 (1.21, 1.49) | 1.24 (0.36, 4.24) |
| **Wave** |  |  |  |  |
| **Initial** | Reference | Reference | Reference | Reference |
| **Alpha** | 1.20 (0.37, 3.90) | 1.24 (0.35, 4.48) | 1.60 (0.35, 7.31) | 5.22 (0.24, 110.2) |
| **Delta** | 0.57 (0.21, 1.50) | 0.64 (0.23, 1.79) | 0.65 (0.17, 2.48) | 0.19 (0.01, 3.37) |
| **Omicron** | 0.37 (0.15, 0.90) | 0.39 (1.15, 1.07) | 0.32 (0.09, 1.16) | 0.12 (0.01, 1.77) |
| **Number of initial symptoms (per symptom)** | 1.30 (1.20, 1.40) | 1.25 (1.16, 1.35) | 1.34 (1.21, 1.49) | 1.79 (1.34, 2.38) |
| **Myocardial Infarction** | 0.57 (0.23, 1.39) | 0.50 (0.18, 1.36) | 1.19 (0.32, 4.44) | 0.18 (1.34, 2.38) |
| **Stroke** | 1.46 (0.47, 4.52) | 1.09 (0.34, 3.53) | 5.24 (0.76, 38.3) | 5.39 (0.20, 143.6) |
| **Atrial Fibrillation** | 0.61 (0.28, 1.31) | 0.57 (0.25, 1.31) | 0.34 (0.12, 0.94) | 1.66 (0.20, 13.9) |
| **Sleep apnea** | 1.14 (0.68, 1.89) | 1.11 (0.64, 1.92) | 0.94 (0.48, 1.85) | 0.79 (0.19, 3.32) |
| **COPD or Asthma** | 0.89 (0.17, 4.80) | 1.50 (0.23, 10.0) | 0.25 (0.03, 2.27) | 1.44 (0.03, 72.5) |
| **Immunodeficiency** | 0.57 (0.26, 1.28) | 0.56 (0.25, 1.27) | 0.44 (0.16, 1.16) | 0.70 (0.10, 5.21) |
| **Vaccination before COVID-19** | 0.81 (0.44, 1.49) | 0.96 (0.50, 1.84) | 0.97 (0.39, 2.42) | 0.50 (0.08, 3.33) |
| **Vaccination after COVID-19** | 1.57 (0.60, 4.13) | 2.09 (0.75, 5.84) | 1.12 (0.31, 4.07) | 0.11 (0.01, 1.62) |
| **Hispanic ethnicity** | 1.73 (0.95, 3.14) | 1.83 (0.99, 3.39) | 1.63 (0.72, 3.72) | 0.96 (0.08, 10.95) |
| **Subjective socioeconomic status (per unit higher)** | 0.90 (0.79, 1.03) | 0.92 (0.80, 1.06) | 0.88 (0.73, 1.07) | 0.88 (0.59, 1.31) |
| **Highest education** |  |  |  |  |
| **No high school** | N/A | N/A | N/A | N/A |
| **High school graduate** | 0.69 (0.08, 6.03) | 0.70 (0.08, 5.92) | 0.53 (0.04, 6.70) | N/A |
| **College degree** | 1.17 (0.23, 5.97) | 1.16 (0.23, 5.72) | 0.63 (0.10, 3.90) | N/A |
| **Graduate degree** | 0.99 (0.19, 5.12) | 0.97 (0.19, 4.88) | 0.50 (0.08, 3.16) | N/A |
| **Healthcare worker** | 1.02 (0.98, 1.07) | 0.86 (0.52, 1.41) | 0.93 (0.48, 1.83) | N/A |
| **Pre-COVID-19 Depressive Symptoms** | 1.08 (1.01, 1.16) | 1.09 (1.02, 1.18) | 1.12 (1.03, 1.22) | 1.20 (0.98, 1.48) |
| **Pre-COVID-19 Anxiety Symptoms** | 1.04 (0.97, 1.12) | 1.04 (0.97-1.12) | 1.03 (0.94, 1.13) | 0.92 (0.74, 1.13) |
| **Pre-COVID-19 financial insecurity** | 1.64 (1.02, 2.63) | 1.61 (0.98-2.64) | 1.75 (0.93, 3.28) | 4.58 (1.24, 16.83) |

eTable 3 Legend: Sensitivity analyses with only Model 3 shown for each sensitivity analysis compared to the primary analysis (column 1) for only those with pre-COVID baseline survey (column 2), only persistent symptoms at the time of survey completion (column 3), and only severe symptoms (column 4). Overall results across the sensitivity analyses were similar with number of symptoms during acute infection remaining highly significant, and similar trends across variant waves. One notable exception is that female sex was associated with persistent symptoms. Abbreviations: OR = Odds Ratio, CI = Confidence Interval, COPD=Chronic Obstructive Pulmonary Disease
